# Debt, health and HIV spending after the COVID-19 crisis: Assessing alternatives for relief mechanisms and additional financing through a cross-country macroeconomic study

**DOI:** 10.1101/2022.10.04.22280691

**Authors:** Edward Dyson, Alexandra Murray-Zmijewski, Jose-Antonio Izazola-Licea, Tomas Lievens

**Author notes:** These authors contributed equally to this work (Edward Dyson, Alexandra Murray-Zmijewski). These authors also contributed equally to this work.

## Abstract

This study addresses the following policy questions: What forms of debt relief and additional financing are most effective in mitigating the fiscal impact of the COVID-19 pandemic in countries that are at risk of or facing debt distress? What would the subsequent impact be on government health and HIV financing up until 2030? To answer these questions, five debt relief and additional financing options are hypothetically applied to seven Sub-Saharan African countries using a macro-fiscal programming framework and debt scenario modelling. Aggregate impacts demonstrate that, on average, the COVID-19 period has had a significant impact on government health and HIV-related expenditure. Each of the five presented options studied are shown to have an iteratively greater impact in mitigating the effects of the pandemic on government health and HIV-related expenditure. However, none of the debt relief and additional financing options succeed in offsetting the loss of health and HIV fiscal space that has occurred during the COVID-19 pandemic. Adequate solutions to make up for this shortfall require the consideration of options beyond the current policy dialogue.

## Introduction

The COVID-19 pandemic has been devastating for the global economy. COVID-related ‘lockdown’ measures have induced slowdowns in demand, triggering a deep global recession. The subsequent impact on government finances has been severe and is expected to have significant long-term impacts on domestic expenditures, including financing for health and, more specifically, HIV. A part of this fiscal picture is the debt situation. With governments attempting to offset the worst of the pandemic through increased spending to assist with areas such as rising health costs, social security payments, and business support amid plummeting fiscal revenues, an alarming debt situation looms.

In response, the global development community is exploring different kinds of debt relief and additional financing to help low- and middle-income countries (LMICs) expand their budgets for social spending to approximate pre-COVID levels. Different forms of relief and financial assistance produce different outcomes for government health and HIV-related financing, however. There is therefore a need to outline the available mechanisms of debt relief and additional financing and to assess their effectiveness in responding to the needs of low- and middle-income countries. This study addresses the following interrelated policy questions: What forms of debt relief and additional financing are most effective in mitigating the fiscal impact of the COVID-19 pandemic on countries that are at risk of or facing debt distress? What would the subsequent impact be for government health and HIV-related financing up until 2030?

To answer these questions, this macroeconomic study explores how debt is impacting fiscal budgets for government health and HIV in seven Sub-Saharan African countries: Cameroon, the Democratic Republic of the Congo (DRC), Kenya, Lesotho, Mozambique, Uganda, and Zambia. It compares the post-COVID debt situation in each country, estimating how the pandemic will impact government finances more generally, and government health and HIV-related expenditures more specifically, until 2030. This research then models the impact of five debt relief and additional financing options on government health and HIV-related expenditures.

This study is a significant contribution to existing literature since it brings together several strands of research and demonstrates how they relate to one another. There are a range of studies that focus on the economic impact of COVID-19 [1, 2], and several which focus on the impact of COVID-19 on HIV financing specifically [3, 4, 5]. In terms of debt relief, there are several papers which discuss the need for a response to COVID-19 and the adequacy of existing debt responses [6, 7, 8, 9]. By bringing together these different strands of literature and undertaking debt scenario modelling, this paper manages to expose the inaccuracy of the idea that debt alleviation will plug the social spending deficit left by COVID-19.

## Debt Relief and Additional Financing Options

This section outlines different forms of debt relief and additional financing, and assesses their effectiveness in responding to the needs of low- and middle-income countries (LMICs) in Sub-Saharan Africa. This research was conducted to better understand the debt policy landscape and identify options that will be modelled later in the study. The debt relief and additional financing options explored in this section are the Debt Service Suspension Initiative (DSSI), the IMF’s Special Drawing Rights (SDRs), and the High-Indebted Poor Countries (HIPC) Initiative.

### Debt Service Suspension Initiative

In April of 2020, during the onset of the COVID-19 pandemic, the G20 announced a suspension of interest payments on debt until the end of the year. This was later extended to include debt due until the end of 2021. The idea behind the DSSI was to implement a fast-acting measure to bring financial resources to countries to aid their response to the COVID-19 crisis. Theoretically, the DSSI offers all poor countries temporary debt service relief by all creditors, as long as the beneficiary countries commit to using the associated savings to increase their social, health, and/or economic spending to address the COVID-19 crisis.

There are three key problems associated with the DSSI. Firstly, many debtor countries are unwilling to participate, at least partly due to their fears about the impact of the scheme on their sovereign ratings and future access to financial markets [10]. Secondly, the initiative is ultimately only a temporary solution to ongoing debt issues in LMICs. The OECD argues that although the DSSI successfully alleviates immediate liquidity pressures, the policy should be supported by country-by-country analyses of sustainability [11]. Thirdly, and perhaps most crucially, all but one private creditors ha thus far rejected participation [12]. Banks, hedge funds and asset management companies, for example, which in some cases make up more than 40% of countries’ external public debt, have resisted calls to participate in debt postponement. Although the [G20](https://www.theguardian.com/world/g20) framework is intended to incorporate private creditors, it lacks effective mechanisms to enforce their participation. Without it, resources freed up from the efforts of other creditors and new emergency financing provided to fight the impact of Covid-19 will effectively be diverted to subsidise non-participating creditors, creating a moral hazard dilemma for those other creditors and minimizing the positive impact of relief to DSSI participants [13].

If fully implemented, the DSSI would have provided more than $12 billion USD in additional liquidity to the 76 least-developed countries in 2020, and an additional $14 billion USD in 2021. Yet, the DSSI has fallen short of expectations and failed to deliver all of the promised financing, delivering a total of $12.9 billion USD from a total of over $26 billion USD [14]. Furthermore, even if the DSSI is fully implemented, it will not be enough to close Africa’s pandemic-response financing gap [15]. Indeed, UNICEF argues that the amount pales in comparison to the more than one-hundred-billion dollars of debt forgiveness provided under the Heavily Indebted Poor Counties Initiatives [16]. The limited relief offered no cancellation, just a temporary standstill, and no private sector or multilateral participation. Thus, the risks posed to the obtention of additional market-based financing has resulted in only 48 out of 73 eligible countries requesting the relief DSSI would provide.^17^

### Special Drawing Rights

Special Drawing Rights (SDRs) are the reserve assets of the International Monetary Fund (IMF). In an effort to deal with the deep fiscal crisis and the additional needs created by the COVID-19 economic shock, and taking into account the mounting debts, IMF member nations approved an issuance resource injection of $650 billion USD—the largest in the IMF’s history [18]. Allocations to low-income countries amounted to around 3% of this, about $21 billion of the $650 billion total [19]. This is despite the fact that high-income economies had already invested over $17 trillion in their own policies between November 2021 and the inception of the COVID-19 shock.

There have been two key voices suggesting that there should be a reallocation of SDRs to ensure low-income countries received a greater share of the assets. The first is the G7, which in June 2021 endorsed a plan to reallocate $100 billion of new SDRs to poorer countries. The second is South African President Cyril Ramaphosa, who has said that from the total SDR allocation, about one-quarter (approximately $162 billion) should be made available to African countries specifically. Furthermore, he called on rich nations to donate their allotments, rather than merely lending them. At the 2021 Annual meetings a new fund was created to channel SDRs reallocations, and despite these calls, the G20 pressed ahead with the standard allocation according to country quotas.

### Highly Indebted Poor Countries Initiative

The Highly Indebted Poor Countries (HIPC) Initiative was initiated by the IMF and the World Bank in 1996 to alleviate over indebtedness of a selected group of countries through bilateral and multilateral partial debt cancellation. The international community supported this action at least partly because HIPCs would then be resume stronger economic growth and use financing and debts in a sustainable manner in the future. To be considered for the initiative, countries had to be in a situation of measured through several thresholds. Assistance was established as conditional on the national governments of these countries meeting a range of economic management and performance targets and undertaking economic and social reforms.

The Multilateral Debt Relief Initiative (MDRI), was adopted by the IMF in late 2005 to top up the HIPC initiative and allow for deeper multilateral cancellation with compensation and support from donor countries. MDRI calls for the cancellation of 100 percent of the claims of three multilateral institutions— the IMF, the International Development Association (IDA), and the African Development Fund—for all countries that meet HIPC criteria [20]. Combined, the MDRI and HIPC initiatives have provided around USD 99 billion in debt relief [21].

## Materials and Methods

### Country Selection Process

A country selection process yielded this list of seven countries to include in the analysis. Firstly, the scope was restricted to LMICs in Sub-Saharan Africa—a region where HIV and debt distress are prevalent in many countries. Secondly, we zoomed in on those African countries that are part of the 30 countries globally that account for 89% of all new HIV infections [22]. This left a total of 18 countries, which were further narrowed down to seven by considering the availability of high-quality data including health data such as National Health Accounts (NHA), the latest National Aids Spending Assessments (NASA), IMF and World Bank debt data, and participation in debt initiatives such as DSSI and the HIPC Initiative. It was also important to target countries with a range of different experiences to allow findings to be more widely applicable. Countries were therefore selected based on geographical variation, rates of HIV prevalence, and risks of debt distress.

### Macro-fiscal Programming Framework

This analysis uses a financial programming framework to model the macro-fiscal indicators for each country’s economy over time. Government health and HIV-related financing projections are made using these underlying economic indicators, i.e., each model is informed by country-specific economic fundamentals and global economic forecasts that impact government health and HIV-related financing. Empirical evidence shows that economic growth is by far the most important determinant of government health spending, followed by changes in total public spending and the reprioritisation of health spending within government budgets [23]. In most countries, fiscal deficits widened in 2020 as reduced economic output led to reductions in fiscal revenue, and as emergency balance of payment support was insufficient to accommodate the increased demands on government expenditures during a time of reduced domestic revenues.

To model the possible impact of the COVID-19 crisis on government health and HIV-related expenditures up to 2030, country-specific data and projections from IMF and government sources are used for each of the seven countries. Data from both before and after the COVID-19 pandemic is analysed for purposes of comparison.

Although the financial programming framework that has been created to inform this study is based on the latest available data and information, the extent to which available datasets incorporate the impact of COVID-19 differs across countries. Data on the macro-fiscal environment in each country is extracted from the IMF World Economic Outlook (WEO) from April 2022 [24], which incorporates the macroeconomic impact from COVID-19 including 2020, 2021, and 2022 fiscal responses. The October 2019 IMF WEO database containing IMF predictions was used to construct the scenario assuming COVID-19 never happened; this discrepancy in data sources accounts for the difference in 2019 data for the scenarios with COVID and the scenario without COVID in all five figures below.

Debt data was primarily gathered from the IMF’s Debt Sustainability Analysis database [25], along with data collected from individual websites for the DSSI, SDRs, and HIPC initiatives. The latest data from the World Health Organisation and National Health Accounts for the respective countries was used to calculate global health expenditures. For HIV-related expenditures, data was utilised from the UNAIDS HIV Financial Dashboard [26] and the National Aids Spending Assessments for the respective countries. Where readily available, official government websites were also accessed to gain budget and expenditure data for 2020 to 2022 health and HIV-related spending. Data was available from Ministry of Finance websites in Cameroon, Kenya, Lesotho, and Uganda. For the DRC, Mozambique, and Zambia, average changes in the government health expenditures (GHEs) of other countries over the three-year period were used as proxies.

### Debt and Additional Financing Scenario Modelling

Options for debt scenario modelling were selected based on an analysis of the existing debt relief and additional financing mechanisms discussed in the literature review at the outset of this paper. In the post-COVID macro-fiscal scenarios, five debt relief and additional financing options were considered. The methodology behind each option is explained below.

1. **DSSI** – The World Bank DSSI website lists the *expected* savings for each eligible country as a proportion of GDP in millions of USD over the period from May 2020 to December 2021 [27]. However, recent IMF Article IV reports include more up-to-date information on the *actual* DSSI arrangements agreed (as in the cases of Cameroon, Kenya, Lesotho, and Uganda). These reports are utilised where available. The estimates or actuals for DSSI are employed to reduce interest payments in 2020 and 2021 for each of the seven countries.
2. **Private sector participation in DSSI** – The World Bank DSSI website also provides a database for country debt by source. The proportion of private debt is calculated, and the same share of DSSI to official multilateral and bilateral debt relief is applied to private debt. These estimates are then used to reduce interest payments in 2020 and 2021. It is assumed that the private sector participate in equal terms to bilateral creditors.
3. **SDR reallocation of $100 billion to low- and middle-income countries (LMICs)** – The IMF website lists each country’s SDR quota based on its number of SDRs as a percentage of the global total [28]. The quotas are used to calculate share amounts from the newly allocated $650 billion USD worth of SDRs (as of August 2021). This amounts to relatively little for each of the countries analyzed here. Therefore, shares are then increased along the lines of the G7 recommendation to provide $100 billion of this allocation to all low- and middle-income countries [29]. The World Bank Development Indicators database was used to find the total GDP of all low- and middle-income countries and then find the proportion of the $100 billion USD worth of SDRs for each of the seven countries selected. This amount was used to offset interest payments over four years (2021 to 2024). Whilst this is done for modelling reasons, it should be noted that the intention of the immediate reallocation would be to provide resources for the early health and economic recovery.
4. **SDR reallocation of $162.5 billion to African countries** – The President of South Africa, Cyril Ramaphosa, recommended that a quarter of the $650 billion USD worth of SDRs should be shared amongst African countries [30]. The same methodology and data sources were used as per the issuance of SDRs outlined in the $100 billion SDR reallocation, the total GDP of Africa was used to determine the share applicable to each of the seven countries. Again, this proportion was offset against interest payments over four years (2021 to 2024). As above, the intention of the immediate reallocation would be to provide resources for the early health and economic recovery.
5. **HIPC Initiative-style debt forgiveness** – The proportion of official multilateral and bilateral debt in DSSI datasets (as per the aforementioned DSSI and DSSI private sector participation) was calculated as a proportion of GDP for each country. This share of GDP was offset against debt over eight years, from 2023 to 2030, which accounts for a lag of several years to allow for the final deal to be negotiated.

To answer our study questions, these five debt relief and additional financing options were compared using scenarios assuming without COVID and with COVID scenarios.

## Results

This section outlines the results of our macro-fiscal programming and debt scenario modelling exercise for the seven countries in question. Firstly, we outline the aggregate results, which demonstrate the impact of COVID-19 on Gross Domestic Product (GDP) and government health and HIV-related spending. Secondly, we model the five debt and additional financing scenarios based on the extent to which they mitigate the damage to finances caused by COVID-19.

### Aggregate Impacts of the COVID-19 period

In five of the seven country case studies, GDP per capita has declined since the onset of the COVID-19 pandemic. This decline in GDP per capita has had a significant knock-on effect on government spending, with reduced tax revenues curtailing the ability of governments to spend. At the same time, there are significant short-term needs for emergency health financing to ensure that countries are well equipped to respond to the pandemic.

All countries require an increased need for health resources as a result of the pandemic, and the pandemic has increased resource needs to achieve HIV/AIDS-related targets including those set by United Nations Sustainable Development Goal (SDG) 3.3.1, which aims to end AIDS as a public health threat by 2030. Despite this, government health expenditure (GHE) is not predicted to increase over the period in six of the seven countries in response to COVID-19. In fact, GHE is predicted to be significantly lower in three of the seven (Kenya, Lesotho, and Mozambique).

In terms of government expenditure on HIV/AIDS (GAE), it is not predicted to increase in response to COVID-19 in six of the seven countries and is predicted to be significantly lower in four of these (Kenya, Lesotho, Uganda and Zambia). This is particularly concerning given that, as of July 2021, UNAIDS predicted that the resource needs of countries in Southern and Eastern Africa to achieve AIDS-related targets are $15.90 per capita, whereas in West and Central Africa they are $4.10 [31].

The impact of COVID-19 on the finances of specific countries is outlined in the figures below. These include the following key findings:

- The average annual loss in GDP per capita up to 2030 is $110 USD. The largest projected loss was in Kenya, estimated at $605 USD per year per capita on average. The DRC and Zambia were the least impacted, with GDP per capita projected to be higher since the onset of the pandemic.
- The average annual rise in the debt-to-GDP ratio is 8%, ranging from 0% in Cameroon to 36% in Zambia. Each of the seven countries is projected to have an increase in their debt-to-GDP ratios, apart from Mozambique which has a tighter fiscal programme scheduled in the latest IMF WEO to pay off debt.
- The average annual loss in GHE per capita up to 2030 is $3 USD. The largest projected average annual loss of GHE per capita up to 2030 is in Kenya at $20 USD. GHE increased in Cameroon and Uganda in response to COVID-19 but is only expected to be maintained in Uganda.
- The average annual loss in GAE per capita up to 2030 is $1.1 USD. The largest average annual projected loss in GAE per capita up to 2030 is in Kenya and Lesotho, with $3 USD each. This has significantly contributed to the increase in resource needs in those countries for achieving SDG 3.3.1 as well as other AIDS-related goals.

### The Impact of Each Debt Scenario

In order to address the shortfall in government health and HIV-related spending outlined above, debt relief options can provide budgetary room to allocate resources to health and HIV. Using the post-COVID macro-fiscal framework, we modelled the five different debt relief options outlined above.

In summary, the average projected impact for the seven countries for each of the five options is as follows:

- DSSI for official multilateral and bilateral debt provides $226 million USD each year over two years.
- Private sector participation for DSSI provides $86 million USD over two years.
- The $100 billion USD SDR reallocation for low- and middle-income countries provides $118 million USD each year over four years.
- The $162.5 billion USD SDR reallocation to Africa provides $713 million USD over four years.
- HIPC Initiative-style debt forgiveness provides $1.7 billion each year over eight years, presumably beginning in 2023. The results for each country-specific case are set out in Table 2

**Table 1:**
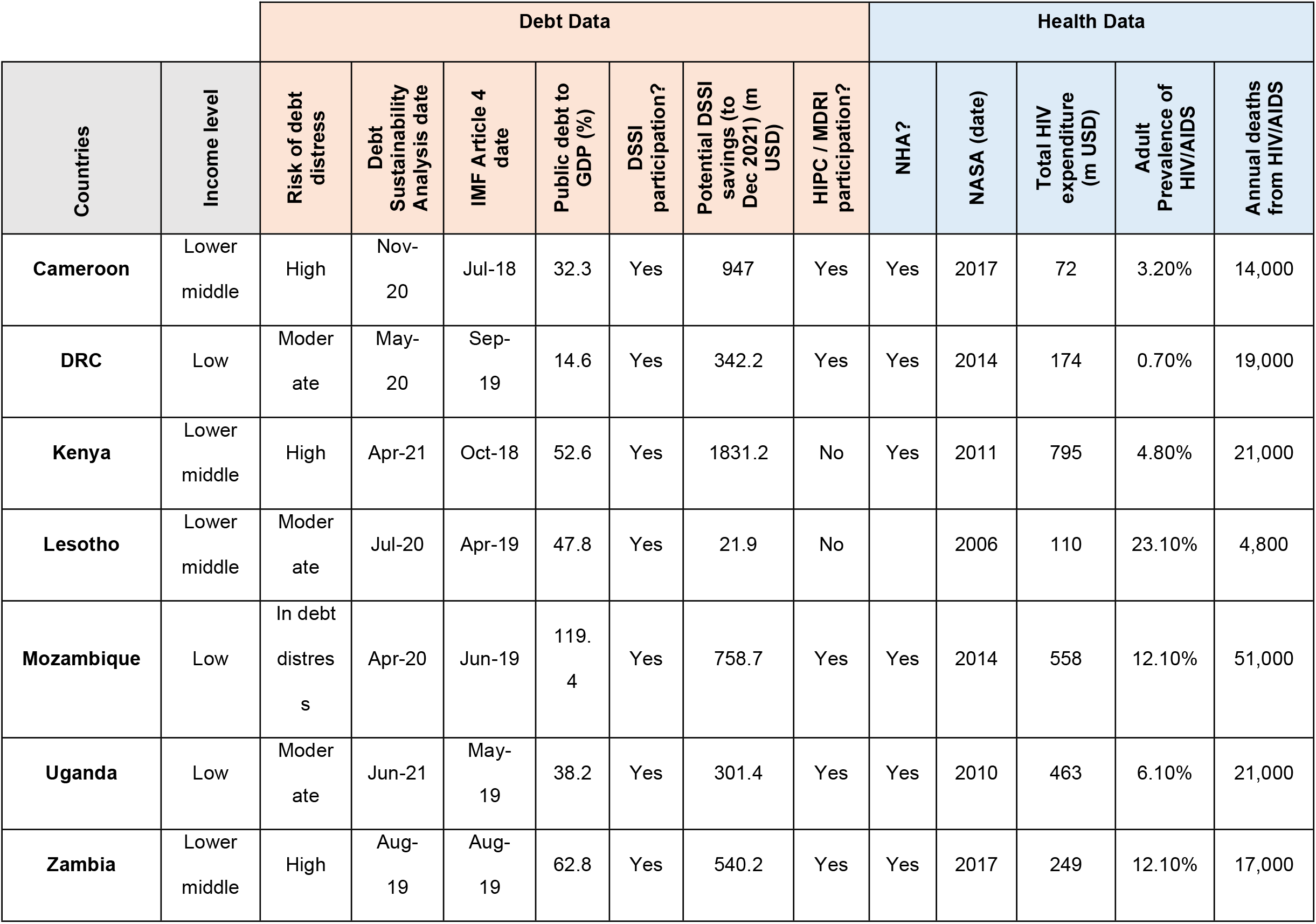
Indicators used to inform the country selection process.

**Table 2:** Potential Fiscal Space from Debt Relief and Additional Financing Options (Million USD) [36, 37, 38, 39].

|  | 2020 | 2021 | 2022 | 2023 | 2024 | 2025 | 2026 | 2027 | 2028 | 2029 | 2030 |
| --- | --- | --- | --- | --- | --- | --- | --- | --- | --- | --- | --- |
| **Fiscal Space Available from DSSI** |  |  |  |  |  |  |  |  |  |  |  |
| Cameroon | 0.0 | 299.5 | 0.0 | 0.0 |  |  |  |  |  |  |  |
| DRC | 0.0 | 342.2 | 0.0 | 0.0 |  |  |  |  |  |  |  |
| Kenya | 380.0 | 723.0 | 268.0 | 252.0 |  |  |  |  |  |  |  |
| Lesotho | 0.0 | 6.0 | 0.0 | 0.0 |  |  |  |  |  |  |  |
| Mozambique | 379.4 | 379.4 | 0.0 | 0.0 |  |  |  |  |  |  |  |
| Uganda | 121.0 | 0.0 | 0.0 | 0.0 |  |  |  |  |  |  |  |
| Zambia | 0.0 | 540.0 | 0.0 | 0.0 |  |  |  |  |  |  |  |
| **Average** | **125.8** | **327.2** | **38.3** | **36.0** |  |  |  |  |  |  |  |
| **Fiscal Space Available from private sector participation in DSSI** |  |  |  |  |  |  |  |  |  |  |  |
| Cameroon | 0.0 | 32.1 | 0.0 | 0.0 |  |  |  |  |  |  |  |
| **DRC** | 0.0 | 6.1 | 0.0 | 0.0 |  |  |  |  |  |  |  |
| **Kenya** | 132.1 | 298.2 | 222.7 | 223.0 |  |  |  |  |  |  |  |
| **Lesotho** | 0.0 | 0.0 | 0.0 | 0.0 |  |  |  |  |  |  |  |
| **Mozambique** | 63.1 | 38.9 | 0.0 | 0.0 |  |  |  |  |  |  |  |
| **Uganda** | 0.0 | 19.1 | 0.0 | 0.0 |  |  |  |  |  |  |  |
| **Zambia** | 0.0 | 618.2 | 0.0 | 0.0 |  |  |  |  |  |  |  |
| **Average** | 27.9 | 144.7 | 31.8 | 31.9 |  |  |  |  |  |  |  |
| **Fiscal Space Available from a \$100 billion SDR reallocation to LMICs** | | | | | | | | | | | |
| **Cameroon** |  | 126.3 | 126.3 | 126.3 | 126.3 |  |  |  |  |  |  |
| **DRC** |  | 158.3 | 158.3 | 158.3 | 158.3 |  |  |  |  |  |  |
| **Kenya** |  | 313.8 | 313.8 | 313.8 | 313.8 |  |  |  |  |  |  |
| **Lesotho** |  | 5.9 | 5.9 | 5.9 | 5.9 |  |  |  |  |  |  |
| **Mozambique** |  | 44.5 | 44.5 | 44.5 | 44.5 |  |  |  |  |  |  |
| **Uganda** |  | 118.6 | 118.6 | 118.6 | 118.6 |  |  |  |  |  |  |
| **Zambia** |  | 61.3 | 61.3 | 61.3 | 61.3 |  |  |  |  |  |  |
| **Average** |  | 118.4 | 118.4 | 118.4 | 118.4 |  |  |  |  |  |  |
| **Fiscal Space Available from a \$162.5 billion SDR reallocation to African countries** | | | | | | | | | | | |
| **Cameroon** |  | 844 | 408 | 367 | 395 |  |  |  |  |  |  |
| **DRC** |  | 470 | 273 | 316 | 361 |  |  |  |  |  |  |
| **Kenya** |  | 2,382 | 2,382 | 2,382 | 2,382 |  |  |  |  |  |  |
| **Lesotho** |  | 44 | 44 | 44 | 44 |  |  |  |  |  |  |
| **Mozambique** |  | 338 | 338 | 338 | 338 |  |  |  |  |  |  |
| **Uganda** |  | 901 | 901 | 901 | 901 |  |  |  |  |  |  |
| **Zambia** |  | 466 | 466 | 466 | 466 |  |  |  |  |  |  |
| **Average** |  | 778 | 687 | 688 | 698 |  |  |  |  |  |  |
| **Fiscal Space Available from HIPC Initiative-style debt forgiveness** |  |  |  |  |  |  |  |  |  |  |  |
| **Cameroon** |  |  |  | 1,524 | 1,642 | 1,774 | 1,912 | 2,054 | 2,188 | 2,326 | 2,465 |
| **DRC** |  |  |  | 161 | 176 | 193 | 210 | 230 | 264 | 293 | 322 |
| **Kenya** |  |  |  | 4,595 | 4,887 | 5,244 | 5,626 | 5,984 | 6,547 | 7,116 | 7,685 |
| **Lesotho** |  |  |  | 44 | 47 | 51 | 53 | 56 | 59 | 63 | 68 |
| **Mozambique** |  |  |  | 393 | 433 | 462 | 534 | 615 | 718 | 819 | 900 |
| **Uganda** |  |  |  | 530 | 574 | 630 | 696 | 759 | 846 | 938 | 1,031 |
| **Zambia** |  |  |  | 1,590 | 1,714 | 1,836 | 1,952 | 2,086 | 2,294 | 2,494 | 2,681 |
| **Average** |  |  |  | 1,262 | 1,353 | 1,456 | 1,569 | 1,683 | 1,845 | 2,007 | 2,165 |

These values can be translated into additional fiscal space for the budgets of each country. If we take these values and allocate them to the health and HIV sectors—as per their current share of gross government expenditure (GGE)—the results are shown in

Figure 3 and Figure 4. The resultant additional fiscal space for health and HIV is projected as follows

**Figure 1:**
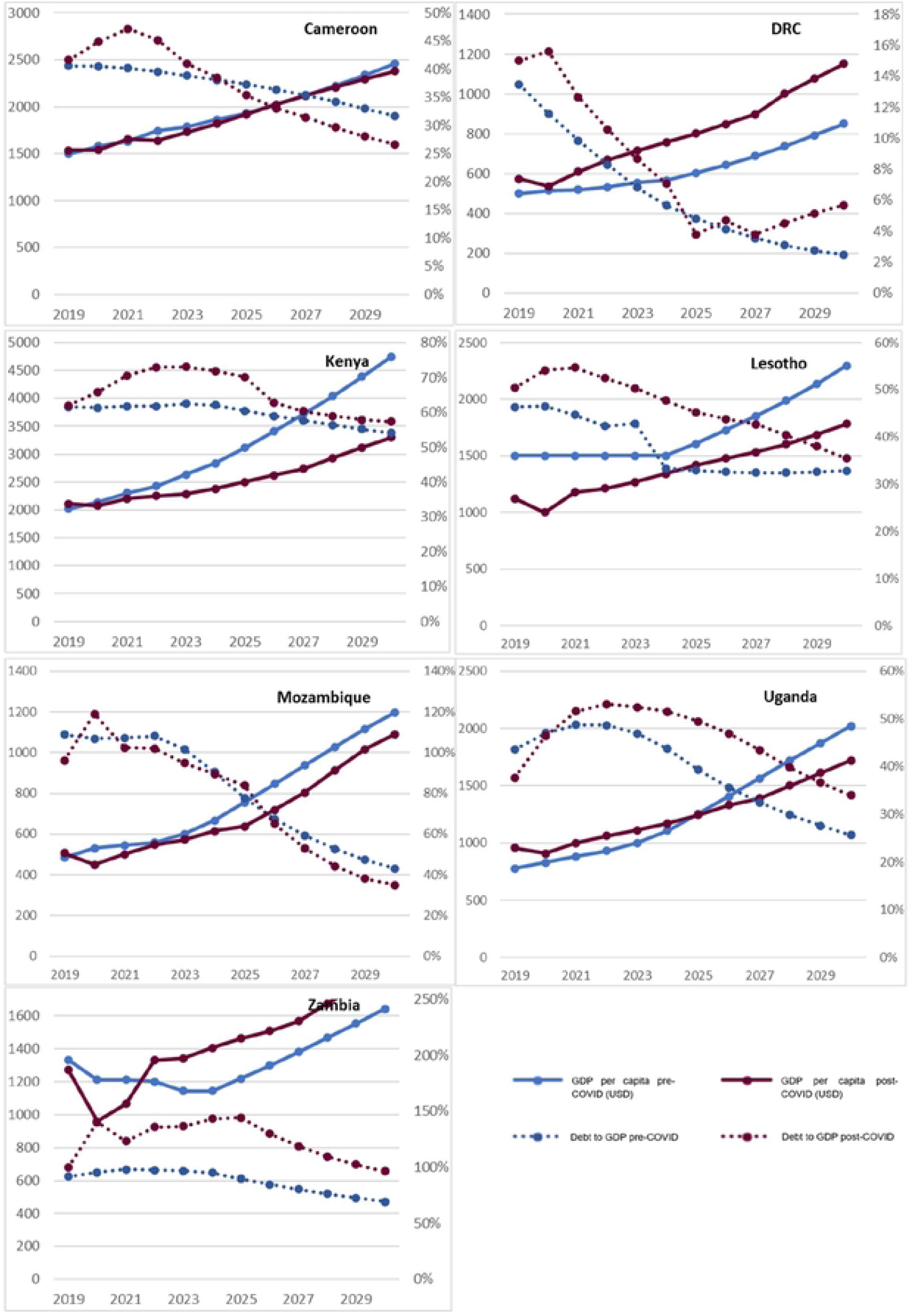
Projections that do and do not include the impact of COVID-19 on GDP per capita (USD) and Debt-to-GDP Ratios [32, 33].

**Figure 2:**
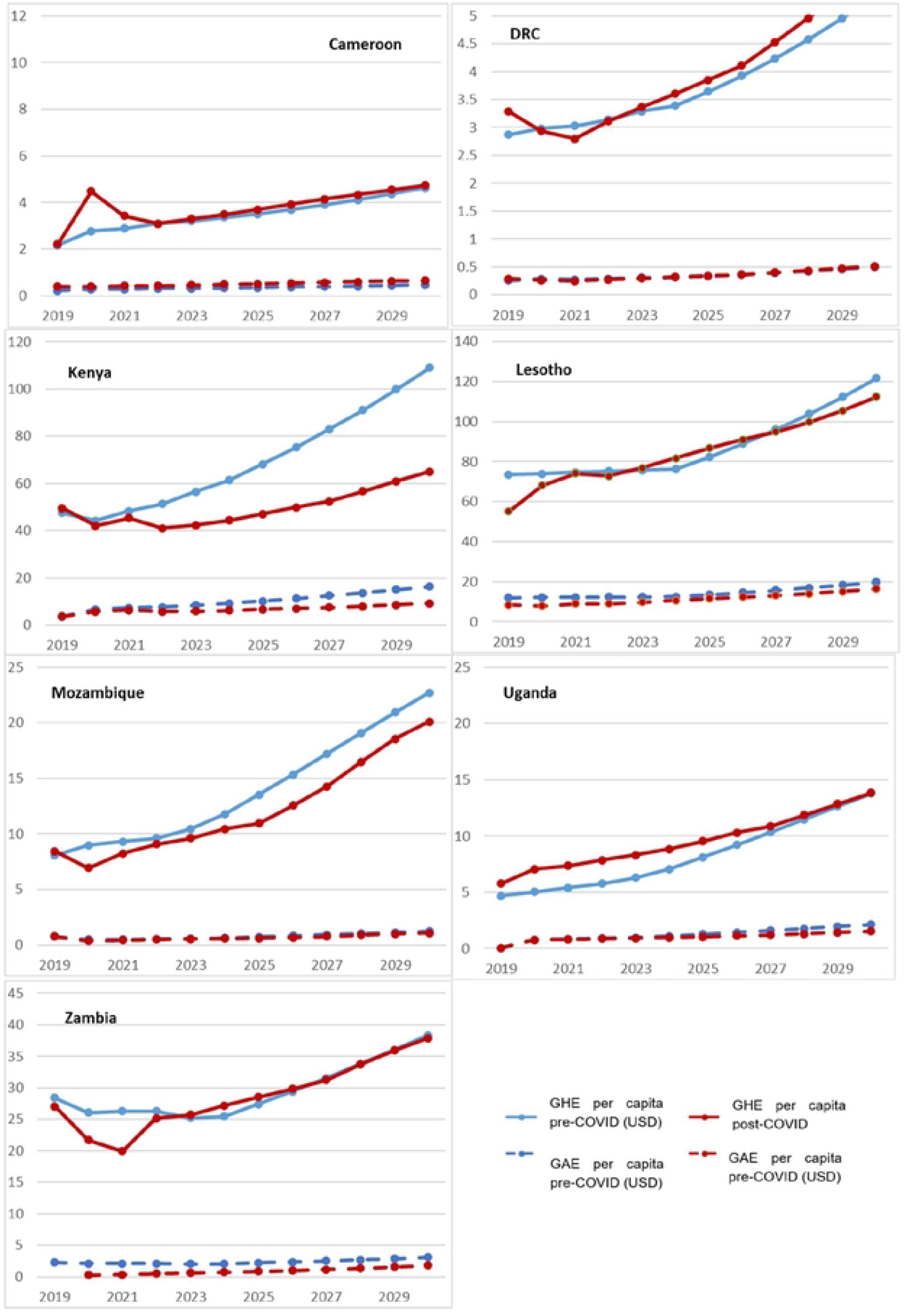
Projections that do and do not include the impact of COVID-19 on GHE and GAE per capita (USD) [34, 35].

**Figure 3:**
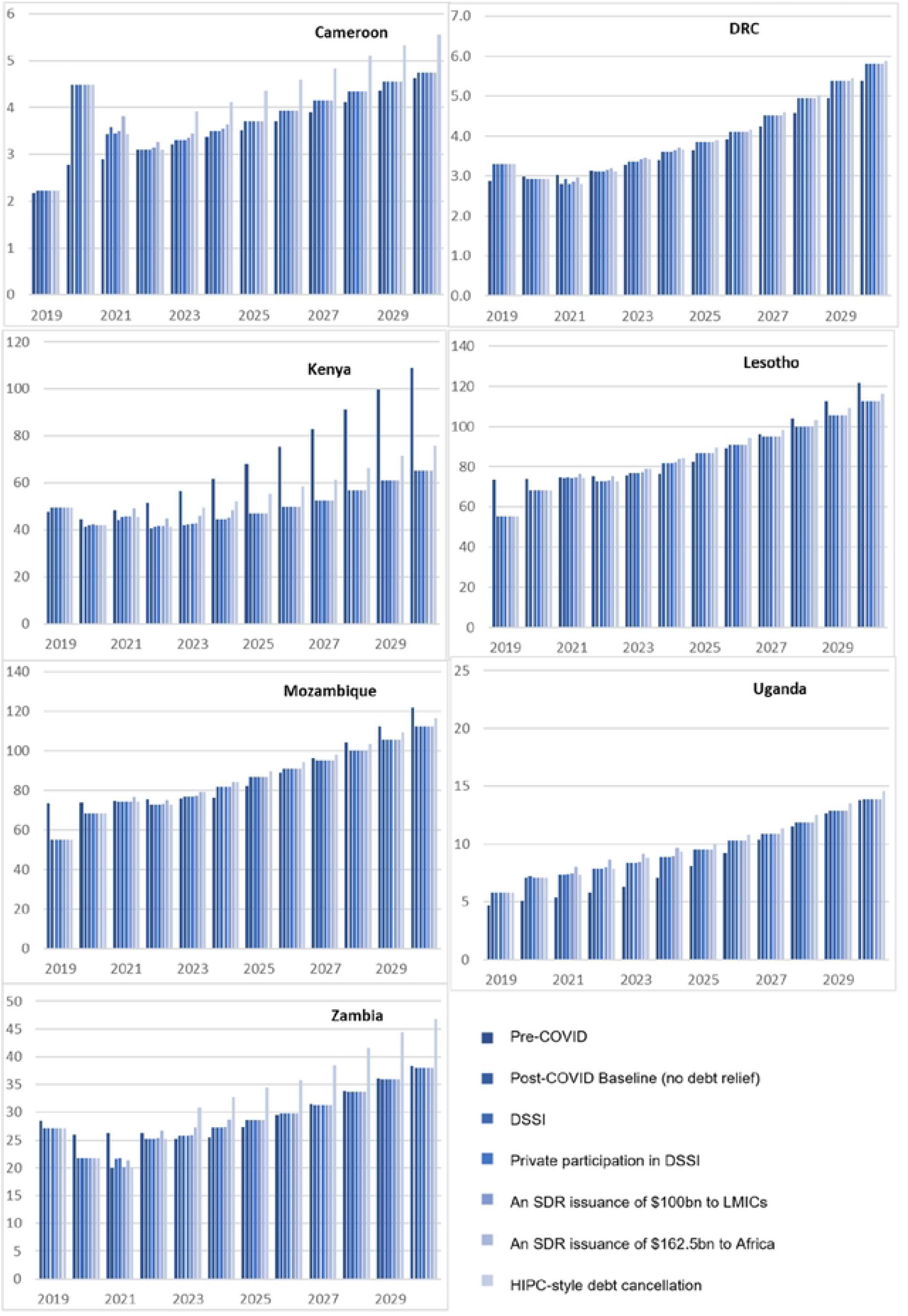
Potential for increase in GHE per capita from Debt Relief and Additional Financing Options (USD)

**Figure 4:**
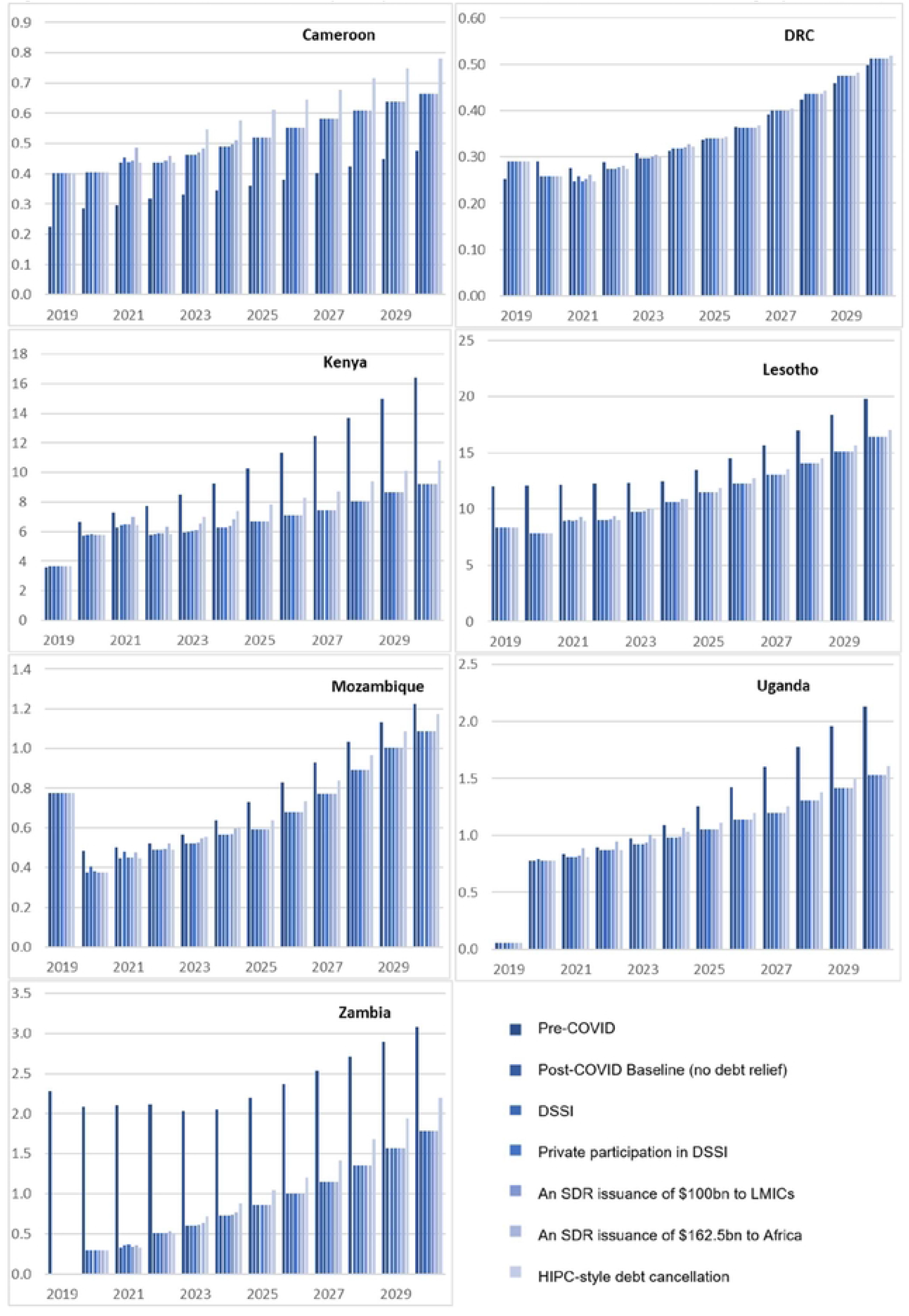
Potential for increase in GAE per capita from Debt Relief and Additional Financing Options (USD)

- **DSSI** – The involvement of donor country lenders in DSSI is projected to save the seven countries $226 million USD each year on average, over two years. This is the equivalent of $7 USD per person in these countries for governments to spend. This translates to an additional annual $0.4 USD per person for health, and an additional $0.03 USD for HIV.
- **Private sector participation in DSSI** – If the private sector joined multilateral lenders in suspending debt servicing repayments, this could save an additional $86 million USD each year over two years for each country on average. This is the equivalent of $5 USD per person in these countries for governments to spend. This could mean an additional $0.3 USD per person for health, and an additional $0.03 USD for HIV.
- **$100 billion SDR reallocation to LMICs** – If $100 billion USD of the SDR quotas were shared with low- and middle-income countries, these seven countries would gain an additional $118 million USD each year for four years for each country on average. This is the equivalent of $4 USD per person in these countries for governments to spend. This could mean an additional $0.3 USD per person for health, and an additional $0.03 USD for HIV.
- **$162.5 billion SDR reallocation to African countries** – If a quarter of the SDR quotas were shared with African countries, these seven countries would gain an additional $713 million USD each year for four years for each country on average. This is the equivalent of $21 USD per person in these countries for governments to spend. This could mean an additional $1.4 USD per person for health, and an additional $0.15 USD for HIV.
- **HIPC Initiative-style debt forgiveness** – If debt relief was provided in a HIPC Initiative style, the countries would receive an additional $1.7 billion USD of fiscal space per year for eight years. This is the equivalent of $45 USD per person in these countries for governments to spend. This could mean an additional $3.1 USD per person for health, and an additional $0.31 USD for HIV.

## Discussion

These aggregate impacts demonstrate that the COVID-19 period has had a significant impact on GDP, GHE, and GAE per capita in several of the countries examined, as well as debt-to-GDP ratios. However, significant disparities should be noted between countries. For example, across the indicators modelled, Cameroon and the DRC are predicted to not be as adversely affected as Lesotho and Kenya. In terms of the potential efficacy of debt relief and additional financing options in mitigating the impact of the COVID-19 period on government health and HIV-related expenditure, each option is shown to have an iteratively greater impact on available budgetary room.

Despite this, it is clear that none of these debt relief options demonstrate sufficient impact to offset the loss of fiscal space for health that has occurred during the COVID-19 period in three of the countries of the study: Kenya, Lesotho and Mozambique. In terms of HIV/AIDS, this is true of five of the seven: Kenya, Lesotho, Mozambique, Uganda and Zambia.

Whilst a full application of DSSI, including multilateral actors, shows a positive impact overall in the medium term, and the involvement of the private sector would contribute further to this, these debt relief and additional financing options subsequently offer minimal amounts of additional fiscal space that can be allocated to health and HIV when compared with the overall loss of fiscal space. We can conclude therefore that the DSSI initiative is bringing very limited results and has proven to be poorly prepared to help countries cope with the economic and health impacts of COVID-19. The fact that the initiative only postponed payments and provided not cancellation make our conclusions more troubling. When the postponed debts are added to the repayment calendar, the actual fiscal pressure will have only grown. The SDR reallocation of the 2021 historical IMF issuance also adds some fiscal space for each country, particularly when we model the Ramaphosa proposal regarding the reallocation of a quarter of SDRs to African countries, but this is still relatively insignificant compared to the amount of budgetary flexibility governments are projected to have had for health and HIV-related spending prior to the pandemic. Finally, the largest amount of impact of all the options is provided by the HIPC Initiative debt relief. This entails actual debt cancellation, rather than temporary relief. This goes much further in reducing the financing gap between scenarios with and without the impact of COVID.

These results have significant implications for the health and HIV outcomes of Sub-Saharan African countries, particularly with regards to achieving SDG 3.3 along with a host of other targets within the SDG framework. In terms of debt relief and additional financing options, the policy debate has so far focused on ensuring that all of DSSI is applied to all of the debt and calls that have been made to include the private sector in such arrangements. It has also included calls for a reallocation of SDRs to low- and middle-income countries. However, it is clear from the results of this study that existing demands and initiatives are relatively marginal, particularly for worst affected countries. The fact that the DSSI is now coming to an end with no further extension, as decided by the G20 in October 2021, leaves tens of countries with high vulnerability and a tricky path to receive such relief. Although every little bit of relief helps, these initiatives do not significantly contribute to mitigating the impact of the COVID-19 pandemic on government health and HIV-related expenditure, and outcomes, in several countries under debt distress.

However, HIPC Initiative-style debt cancellation demonstrates the greatest impact on the long-term prospects for health and HIV in these seven countries up to 2030. Considering historic examples such as the HIPC Initiative helps broaden the debate beyond what is currently being discussed, and can help policymakers realise what is possible with adequate political will. Although it still would fail to meet the fiscal space predicted in the pre-COVID scenario in three countries for health spending and five for HIV/AIDS, the HIPC Initiative debt scenario demonstrates that such bold thinking is necessary to ensure governments are able to offset the worst of the pandemic expenditures and maintain their fiscal space for domestic expenditure on health and HIV. However, the fact that HIPC took between six and nine years to complete means that a much faster and more profound debt restructuring and cancellation scheme must be put in place. The role of private sector needs to be clarified, as well as that of multilaterals, since all actors must participate if true relief is to be provided.

As importantly, the seven country case studies have demonstrated the differential impact of the COVID-19 period and different debt scenarios on a range of Sub-Saharan African countries. For example, the DRC’s post-COVID GDP per capita is projected to be higher than its pre-COVID projection, whereas Kenya is projected to lose $605 USD per capita per year up to 2030. Furthermore, debt cancellation has a significant impact on fiscal space for health for Mozambique, but less so for Zambia’s, whereas the reverse is true for HIV. The differential impact between the seven country case studies also has significant policy implications, since it demonstrates that a blanket approach for all highly indebted Sub-Saharan African countries would be an inefficient use of resources.

### Policy implications

To reiterate the magnitude and importance of these debt situations for policymakers, here they are again, briefly restated: DSSI provides an average additional $0.4 USD per person for health and $0.03 USD for HIV, private sector participation in DSSI provides an additional $0.3 USD per person for health and $0.03 USD for HIV, the $100 billion SDR reallocation to LMICs provides an additional $0.3 USD per person for health and $0.03 USD for HIV, the $162.5 billion SDR reallocation to African countries provides an additional $1.4 USD per person for health and $0.15 USD for HIV, and the HIPC Initiative debt cancelation provides an additional $3.1 USD per person for health and $0.31 USD for HIV. These figures are mostly insufficient when compared to the average annual loss in GHE per capita of $3 USD and the average annual loss in GAE per capita of $1.1 USD. This is particularly the case in Kenya, where the projected average annual loss of GHE per capita is $20 USD, and the average annual projected loss in GAE per capita is $3 USD in Lesotho and Kenya.

Adequate solutions to this shortfall require the consideration of options beyond current thinking. There is a need to recognise that, while debt service suspension and injections of finance via mechanisms such as SDRs do positively contribute to the fiscal position of governments, this does not make enough of an impact to offset the damage done by the pandemic in many countries at risk or in debt distress. Policymakers must come together to consider options that stray beyond current thinking; more radical and drastic options need to be considered, even beyond the HIPC Initiative. Furthermore, it is important not to only focus on debt to create fiscal space for government spending on health and HIV, but to consider all elements of the fiscal diamond (Official Development Assistance, domestic revenue mobilisation, and reprioritisation and efficiency) to find the necessary finances. Finally, ringfencing mechanisms should be considered to ensure that fiscal space freed up via debt relief is used for social spending and health and HIV specifically.

### Limitations

Firstly, debt incurred throughout the COVID-19 pandemic is not all related to expenditure triggered by COVID-19, and determining the exact nature and causality of debt incurred throughout this period is challenging if not impossible. This paper is therefore not concerned with the question why debt has been incurred, and instead focuses on the impact of rising debt burdens on the fiscal space/budgetary room for government health and HIV-related expenditure. This paper does not attempt to isolate non-COVID and COVID-related borrowing, but instead assesses how debt situations in the post-pandemic world impact expenditures up until 2030.

Secondly, debt relief is likely to have externalities on other variables of fiscal space (ODA, domestic revenues, expenditure efficiency), suggesting that attempts to isolate the impact of debt spending on fiscal space for health and HIV-related expenditure may produce inaccurate results. However, our insights based on reviews of previous debt relief initiatives suggest that the impact of debt relief is likely to have predominantly positive, rather than negative, externalities on the factors affecting the fiscal space for government expenditure on health and HIV. This suggests the positive effects of debt relief in these findings may be underestimated, adding additional weight to our resulting recommendations.

Thirdly, the precise macroeconomic impacts of COVID-19, including how these will vary among countries and from the short to the long term, remains uncertain. The analyses here do not purport to generate precise results up to 2030, but instead should be understood as an opportunity to catch a glimpse of the pandemic’s likely impact on government expenditure on health and HIV. To inform timely and corrective action where needed, actual in-country health and HIV-related expenditure should be monitored on a regular basis.

Finally, there is some level of uncertainty over government health and HIV-related spending data. For example, in the case of the DRC, Mozambique, and Zambia, levels from 2019 to 2021 were estimated as official data was not available. Furthermore, much of the predicted health and HIV spending was derived from government budgets where execution rates have been assumed based on past performance. However, actual execution rates may differ to those of previous years.

## Conclusion

The key message of this study is that the current dialogue around debt relief is insufficient compared with what is needed to safeguard health and HIV outcomes in several of the African countries studied. This makes a key contribution to the field of health financing research in response to the COVID-19 pandemic: while previous studies have focused on the impacts of COVID-19 on government health and HIV-related spending and discussed the effectiveness of various debt relief mechanisms, this study ties various elements together and brings into sharp focus the inadequacy of existing debt relief. It is urgent that policymakers take note of the conclusions of this study to better address the pandemic-related health and HIV financing shortfall and advocate for greater action to create the required fiscal space. In terms of future research, a tailored approach and a case-by-case analysis of debt sustainability is needed given the variability of country contexts. This will ensure that each country is able to reduce the impact of the COVID-19 pandemic and maximize the fiscal space available for health and HIV-related spending to help meet HIV-related SDG goals by 2030. Furthermore, there is a need to extend research beyond the debt relief options in this paper to analyse the potential for more radical debt relief mechanisms and to conduct an institutional analysis of the international debt architecture in light of its response to pandemic-related demands.

## Data Availability

Country-specific datasets are available at the following link: 10.6084/m9.figshare.21253584

## Acknowledgements

The authors would like to thank Jaime Atienza Azcona for being interviewed in preparation for this work and for a comprehensive review of a draft version of the paper.

